# Exploring and mitigating potential bias when genetic instrumental variables are associated with multiple non-exposure traits in Mendelian randomization

**DOI:** 10.1101/19009605

**Authors:** Qian Yang, Eleanor Sanderson, Kate Tilling, M Carolina Borges, Deborah A Lawlor

## Abstract

**Background:** Our aim is to produce guidance on exploring and mitigating possible bias when genetic instrumental variables (IVs) associate with traits other than the exposure of interest in Mendelian randomization (MR) studies.

**Methods:** We use causal diagrams to illustrate scenarios that could result in IVs being related to (non-exposure) traits. We recommend that MR studies explore possible IV-non-exposure associations across a much wider range of traits than is usually the case. Where associations are found, confounding by population stratification should be assessed through adjusting for relevant population structure variables. To distinguish vertical from horizontal pleiotropy we suggest using bidirectional MR between the exposure and non-exposure traits and MR of the effect of the non-exposure traits on the outcome of interest. If vertical pleiotropy is plausible, standard MR methods should be unbiased. If horizontal pleiotropy is plausible, we recommend using multivariable MR to control for observed pleiotropic traits and conducting sensitivity analyses which do not require prior knowledge of specific invalid IVs or pleiotropic paths.

**Results:** We applied our recommendations to an illustrative example of the effect of maternal insomnia on offspring birthweight in the UK Biobank. We found little evidence that unexpected IV-non-exposure associations were driven by population stratification. Three out of six observed non-exposure traits plausibly reflected horizontal pleiotropy. Multivariable MR and sensitivity analyses suggested an inverse association of insomnia with birthweight, but effects were imprecisely estimated in some of these analyses.

**Conclusions:** We provide guidance for MR studies where genetic IVs associate with non-exposure traits.

**Key messages:**

- Genetic variants are increasingly found to associate with more than one social, behavioural or biological trait at genome-wide significance, which is a challenge in Mendelian randomization (MR) studies.
- Four broad scenarios (i.e. population stratification, vertical pleiotropy, horizontal pleiotropy and reverse causality) could result in an IV-non-exposure trait association.
- Population stratification can be assessed through adjusting for population structure with individual data, while two-sample MR studies should check whether the original genome-wide association studies have used robust methods to properly account for it.
- We apply currently available MR methods for discriminating between vertical and horizontal pleiotropy and mitigating against horizontal pleiotropy to an example exploring the effect of maternal insomnia on offspring birthweight.
- Our study highlights the pros and cons of relying more on sensitivity analyses without considering particular pleiotropic paths versus systematically exploring and controlling for potential pleiotropic paths via known characteristics.

## Introduction

Mendelian randomization (MR) is a special case of instrumental variable (IV) analysis where single nucleotide polymorphisms (SNPs) randomly allocated at conception are used as the IVs.^(1, 2)^ MR requires three key assumptions: first, IVs are strongly associated with an exposure of interest (relevance); second, there are no common causes between IVs and an outcome of interest (independence); and third, IVs influence the outcome only through the exposure (exclusion restriction).^(1, 2)^ While the relevance assumption can be tested, the independence and exclusion restriction assumptions are difficult to verify and only their plausibility can be explored.^(3)^ One common approach to date is to test for associations between genetic IVs and a range of non-exposure traits in either one- or two-sample setting as a way to assess the specificity of the genetic IV.^(4-7)^

With increasing sizes of genome-wide association studies (GWAS), and more extensive coverage of the genome due to imputation with more comprehensive panels, SNPs are increasingly found to associate with multiple traits.^(8, 9)^ Therefore, we aim to develop guidance for assessing potential violations of independence and exclusion restriction assumptions when genetic IVs are associated with other (non-exposure) traits. This paper is laid out as follows. In section 1, we use directed acyclic graphs (DAGs) to illustrate four scenarios that could result in an association of a genetic IV with a non-exposure trait and highlight which scenarios would bias MR estimates. In section 2, we describe different methods for discriminating between scenarios and methods for mitigating against potential bias for both one- and two-sample settings. In section 3, we apply this framework to an MR analysis exploring the potential causal relationship between maternal insomnia and offspring birthweight in the UK Biobank (UKB). In section 4, we end with a discussion of our recommendations.

### Scenarios that could explain associations of genetic IVs with multiple traits

There are four broad scenarios consistent with genetic IVs (Z) being associated with multiple traits (Table 1). ***Population stratification*** (PS; DAGs 1.1-1.3 in Table 1), might occurs due to the study including subgroups of people with different ancestry or who were born or live in different geographical locations. If the distribution of SNPs and of non-exposure traits (W) differs by these subgroups, then this PS is a common cause of Z and W and generates an association between them.^(10)^ As an example of this, evidence shows that place of birth in UKB has been associated with genetic IVs for education, height and body mass index (BMI), and also with health outcomes.^(11)^ If PS affects the distribution of Z and the outcome (Y) directly or via W, PS could confound MR estimates (DAGs 1.1-1.2). This would represent a violation of the independence assumption. If PS confounds the Z-exposure (X) association, Z could still be used to estimate the unbiased effect of X on Y if PS did not affect Y independently of X (DAG 1.3).

**Table 1.**
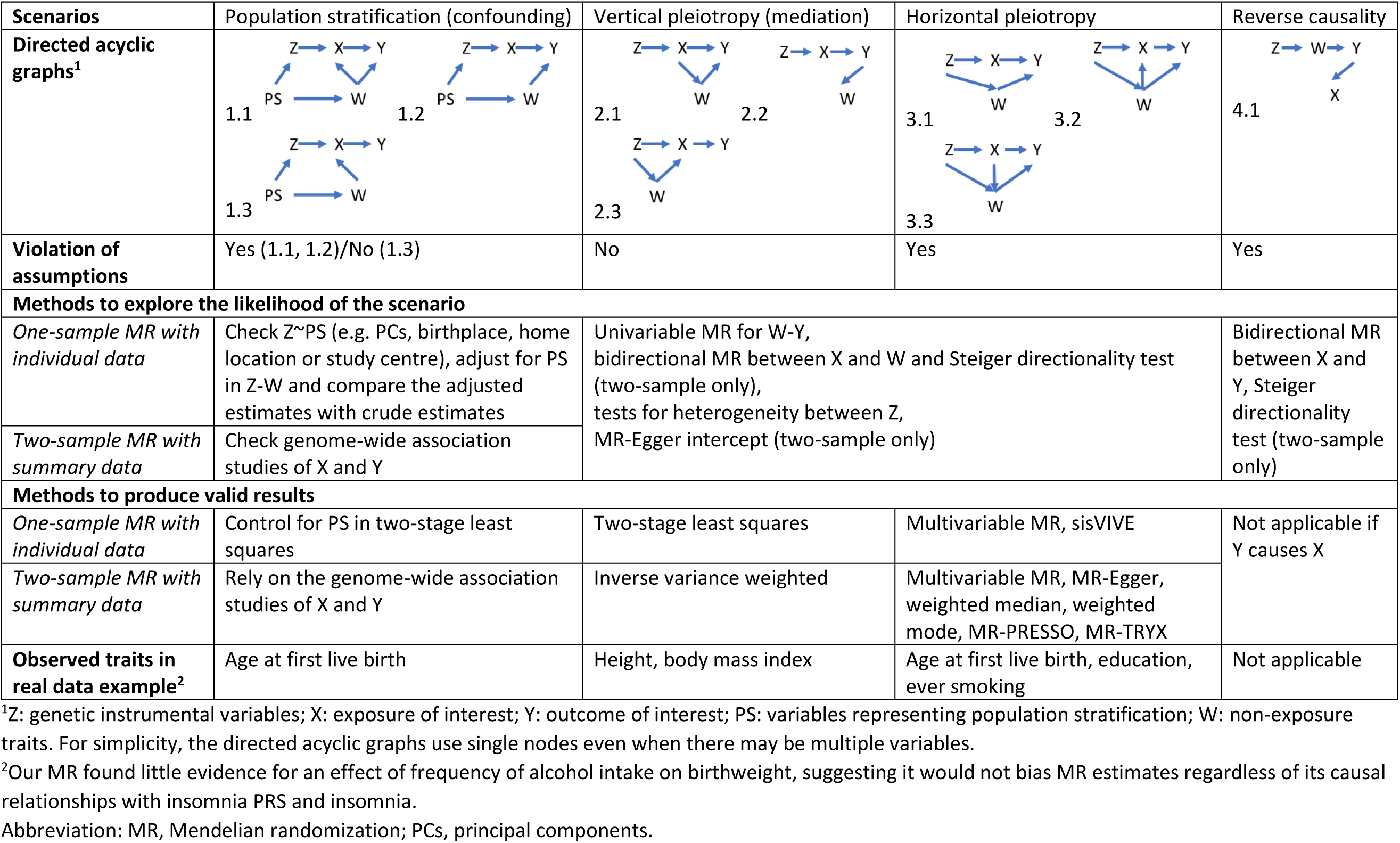
Scenarios when an unexpected IV-non-exposure trait association could be observed.

Pleiotropy refers to the association of a SNP with multiple phenotypes, and has two types: vertical (also known as spurious or false) and horizontal (also known as genuine or true).^(12)^ In the scenario of ***vertical pleiotropy*** (DAGs 2.1-2.3 in Table 1), Z is a cause of X, which in turn affects Y. Despite pleiotropic associations of Z with X and W, the effect of Z on Y is fully mediated by X. Therefore, the exclusion restriction assumption is not violated.^(13)^ In the scenario of ***horizontal pleiotropy*** (DAGs 3.1-3.3 in Table 1), Z is a cause of X and W, and both of them affect Y independently. This violates the exclusion restriction assumption leading to potential bias in MR estimates.^(13)^

In the scenario of ***reverse causality*** (DAG 4.1 in Table 1), Z is really a primary cause of Y, which in turn affects X. As such, inclusion of Z into IVs for X would give a biased X-Y association due to a violation of the exclusion restriction assumption.^(14)^ With respect to the focus of this paper, this would only result in an association of Z with W if Z directly or indirectly influences W (DAG 4.1). We have included this scenario for completeness. However, exploration of Z-W associations is not a good way of identifying causal directions between X and Y. Bidirectional MR and Steiger directionality test should be more suitable for exploring causal directions between any two traits,^(14)^ and will be described in recommendation 4 of the next section.

### Recommendations for exploring above scenarios and to obtain unbiased MR estimates

Having described the different scenarios that could result in genetic IVs relating to non-exposure traits below we provide a list of recommendations for first identifying such non-exposure traits and then exploring which are likely to bias the main MR results and how that might be mitigated (summarised in Table 1).

#### 1. Searching more thoroughly for genetic IV-non-exposure trait associations

To date most MR studies have explored associations of genetic IVs with potential confounders of exposure-outcome associations. By definition exposure-outcome confounders are unlikely to influence genetic variants (which are fixed at conception) and the association of genetic IVs with several non-exposure traits are likely to reflect violation of the exclusion restriction via pleiotropy (i.e. the direction of the arrow will go from genetic variants to the confounders rather than the other way). Consequently, selection of non-exposure trait associations should aim to explore violation of the independence and exclusion restriction assumptions and their specific mechanisms by exploring associations with any risk factors for the outcome, rather than focus solely potential exposure-outcome cofounders.

Once this is acknowledged there are two broad approaches that could be used to identify non-exposure traits that genetic IVs might influence in either one- or two-sample MR. One is to use prior/existing knowledge of the key causes of the outcome and then examine whether the genetic exposure-IV relates to any of these non-exposure causes of the outcome. The second is to undertake a hypothesis free comprehensive genotype-to-phenotype (also known as Phenome-wide) approach, in which we use automated systems to explore all possible associations of our genetic IVs.^(15, 16)^ There are differing pros and cons of these two approaches including different challenges relating to balance between valid application of the following recommendations 3-5 versus a greater reliance on sensitivity analyses in recommendation 6. We explore this further in the discussion.

#### 2. Assessing the impact of population stratification

In one-sample MR with individual participant data, we recommend exploring associations of Z with as many indicators of PS as possible. These could include place (country, region, town, longitude/latitude) of birth and residence, study centre, and genetic principal components of ancestral background. Adjusting the Z-W associations for these sources of PS and exploring whether this alters the association is also useful for exploring PS. If the association was attenuated after adjustment, it would suggest that the Z-W association may be driven by PS. In two-sample MR using summary statistics, the data have typically been generated a priori and thus the investigators are limited in what they can do to account for PS. However, they can still check whether the original GWAS has used robust genomic control methods to properly account for PS. Newly developed two-sample MR methods (MR-PRESSO,^(17)^ GSMR,^(18)^ LCV^(19)^ and GIV^(20)^) may not be able to overcome PS either if PS is not controlled in the original GWAS, as acknowledged by Koellinger *et al*.^(10)^

#### 3. Assessing bias due to horizontal pleiotropy by using MR to explore the W-Y association

After excluding the possibility of bias by population stratification, it is important to investigate whether unexpected Z-W associations might be explained by horizontal pleiotropy. We recommend first undertaking MR to explore whether there is evidence for the effect of W on Y. This requires valid genetic IVs for W, which may not always be available, and sufficient statistical power to precisely estimate the W-Y association. It is also important to consider the strength of the genetic IVs for W, as weak instrument bias would tend to bias the estimate towards the observational association in one-sample MR but to the null in two-sample MR with non-overlapping samples and increase the standard errors of the estimate.^(21)^ If there are valid and strong genetic IVs for W and these provide (convincing) evidence that W does not affect Y, then there cannot be violation of the exclusion restriction criteria via W. If there is evidence for an effect of W on Y, or it is not possible to determine this, then bidirectional MR of the effect of X-W versus W-X is valuable (next point).

#### 4. Distinguishing vertical from horizontal pleiotropy by testing causal directions between X and W

If there is no causal effect between X and W, horizontal pleiotropy (DAG 3.1) would be more plausible than vertical pleiotropy (DAGs 2.1-2.3). If bidirectional MR suggests that X causes W, vertical pleiotropy (DAGs 2.1-2.2) may be more plausible than horizontal pleiotropy (DAGs 3.1-3.2), although we could not fully rule out the possibility that W mediates the effect from Z to Y partly independently of X (DAG 3.3). However, if bidirectional MR suggests that W causes X, W could be a confounder of X and Y and horizontal pleiotropy (DAG 3.2) may be more plausible than vertical pleiotropy (DAGs 2.1-2.2), although we could not fully rule out the possibility that W cannot affect Y independently of X (DAG 2.3).

Bidirectional MR can be conducted in either one- or two-sample setting,^(22, 23)^ but could be misleading when there is marked difference in statistical power between X-W versus W-X associations. For example, if the power for W-X association is low (relative to the power for X-W association) it may appear that there is no causal effect of W on X even in the presence of such an effect. Additionally, overlapping SNPs in the GWAS of X and W can make it unclear which SNPs to select as valid IVs for X and W in bidirectional MR.^(24)^ In two-sample setting, Steiger directionality test can help to identify (independent) valid IVs for X or W by comparing the variance explained by each SNP in X to that in W, as it assumes that a valid IV should explain more variance in trait A than in trait B if trait B is a downstream effect of the trait A.^(25)^ However, Steiger directionality test could be misleading if measurement errors in X and W are substantially different.^(25)^ For example, insomnia is measured much less accurately than height or BMI in UKB (see real data example in the next section).

#### 5. Adjusting for potential horizontal pleiotropic effects via known non-exposure traits

Where there is evidence (from 3 and 4 above) that there may be bias due to horizontal pleiotropy (DAGs 3.1-3.2) from specific non-exposure traits (W), multivariable Mendelian randomization (MVMR) can be used to obtain unbiased estimates in one- and two-sample settings.^(26)^ MVMR requires not only IVX (IV for X)-X and IVW-W associations but also IVX-W and IVW-X associations, which means two-sample MR studies using summary statistics have to access to full results of the original GWAS. If W mediates both Z-Y and X-Y associations (DAG 3.3), controlling for W in MVMR obtains the direct effect rather than the total effect of X on Y, while its total effect should be estimated by using a subset of SNPs only related to X.^(26)^ MVMR can also be used to estimate direct effects of correlated traits on an outcome as long as the genetic IVs independently predict each trait. Limitations of MVMR have been discussed by Sanderson *et al*., e.g. the strengths of IVs may decrease dramatically after attempting to including many non-exposure traits in the estimation.^(26)^

#### 6. Exploring and controlling for bias due to horizontal pleiotropy via unknown traits

It is possible that MVMR adjusting for horizontal pleiotropy via known/measured traits is still biased by unknown/unmeasured traits. Therefore, sensitivity analyses that explore bias due to unbalanced ‘unmeasured’ horizontal pleiotropy will still be required. In one- and two-sample MR, we recommend initial exploration of this by assessing between SNP heterogeneity.^(27)^ This should be done even if SNPs are being combined into a single polygenic risk score (PRS). In one-sample MR heterogeneity is commonly explored by ‘overidentifying’ tests,^(28)^ while in two-sample MR using summary data the Cochran’s Q statistic is an equivalent test.^(27)^ If the exposure causes the outcome and IVs are valid, we expect the effect of the IV on the outcome to be proportional to its effect on the exposure across genetic IVs. Therefore, heterogeneous causal estimates across genetic IVs are indicative of invalid IVs. Most of the sensitivity analyses that have been developed for addressing horizontal pleiotropy aim at exploring the presence or being robust to heterogeneous (potentially invalid) IVs. Table 2 summarises the commonly used methods (i.e. sisVIVE^(29)^ for one-sample MR and MR-Egger,^(30)^ weighted median,^(31)^ weighted mode,^(32)^ MR-PRESSO,^(17)^ MR-TRYX^(33)^ for two-sample MR), including their additional assumptions. It is important to recognise that (i) heterogeneity tests can only be used where there are multiple SNPs, (ii) some methods are statistically inefficient and (iii) most methods have been developed for the two-sample MR setting.

**Table 2.** Summary of select sensitivity analyses for exploring bias due to horizontal pleiotropy in MR.

| Name | Brief description | Assumptions<sup>1</sup> | Other issues |
| --- | --- | --- | --- |
| *For one-sample MR* |  |  |  |
| sisVIVE<sup>(29)</sup> | It is an extension to two-stage least squares, which incorporates LASSO penalization. | At least 50% of IVs are valid. | It works for continuous outcomes only, requires a large amount of computer memory, and the current implementation do not provide 95% CIs. |
| *For two-sample MR* |  |  |  |
| MR-Egger<sup>(30)</sup> | It allows a non-zero intercept to test horizontal pleiotropy. | Instrument strength independent of direct effect | It is sensible to outliers and tends to suffer from low statistical power. |
| Weighted median<sup>(31)</sup> | It is defined as the median of a weighted empirical density function of the Wald ratio estimates. | At least 50% of weight comes from valid IVs. | Nil |
| Weighted mode<sup>(32)</sup> | It calculates the weighted mode of the Wald ratio estimates. It will be unbiased even if the majority of SNPs could be invalid but providing the set of SNPs which form the largest homogeneous cluster are valid.<sup>(27)</sup> | Zero modal pleiotropy assumption | Researchers need to choose a bandwidth to obtain the clustering effect, and different bandwidths might provide inconsistent estimates.<sup>(27)</sup> |
| MR-PRESSO<sup>(17)</sup> | It assesses horizontal pleiotropy based on the contribution of each SNP to heterogeneity and provides adjusted MR estimates by removing outlier SNPs. | Instrument strength independent of direct effect; Outliers (identified via MR-PRESSO global test) are risen due to potential horizontal pleiotropy. | After removing outlier SNPs, the standard errors would decrease. Therefore, it would be more likely to reject the null. |
| MR-TRYX<sup>(33)</sup> | It assesses horizontal pleiotropy based on the contribution of each SNP to heterogeneity and attempts to adjust for their horizontal pleiotropic effects using extra publicly available GWAS from MR-Base. | Outliers (identified via RadialMR<sup>(53)</sup>) are risen due to potential horizontal pleiotropy. | GWAS from MR-Base may not cover the whole genome or conducted in the target population (e.g. only female participants). |

<sup>1</sup>Extra assumptions except for the three key MR assumptions.
Abbreviations: CI, confidence interval; GWAS, genome-wide association studies; IV, instrumental variable; MR, Mendelian randomization; SNP, single nucleotide polymorphisms.

### Real data example

We use the effect of maternal insomnia on offspring birthweight as a motivating example, as it has been suggested that having insomnia and other forms of sleep disturbance may be associated with lower offspring birthweight though results are inconsistent.^(24, 34, 35)^ We explore this question in UK Biobank women,^(36)^ using a PRS that combines 80 genome-wide significant SNPs (listed in Supplementary Data 1) from the largest GWAS of insomnia in women.^(37)^ We tested associations of the PRS with six observed traits (maternal height, BMI, age at first live birth, education, frequency of alcohol intake and ever smoking) that are known to (or could plausibly) influence offspring birthweight, and found that the PRS was associated with all of them (Figure 1). We demonstrate how to use the above recommendations in both one- and two-sample MR analyses, with full details in Supplementary Methods.

**Figure 1.**
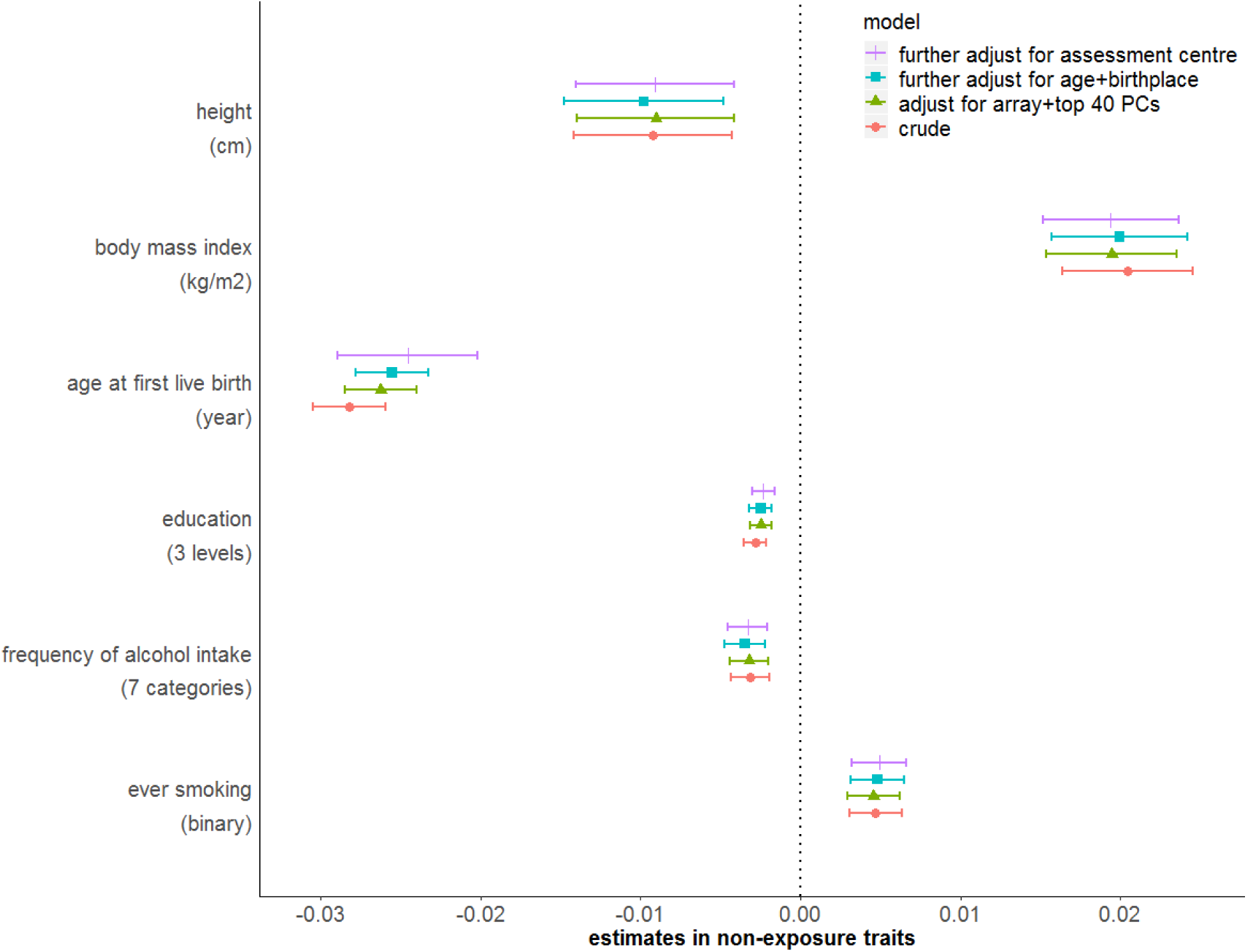
Associations of polygenetic risk score (PRS) for insomnia with six non-exposure traits before and after adjustment for population stratification. Estimates are differences in mean non-exposure traits or log odds ratio (ever smoking) per allele increase in PRS. Supplementary Table 1 summarizes how education, frequency of alcohol intake and ever smoking are coded in this study.

UKB is a cohort of 503,325 men and women who were on the National Health Service registry, aged between 40-69 years and living within 25 miles from one of 22 assessment centres.^(36)^ One-sample MR included genetically unrelated women of European descent who reported frequency of insomnia, had experienced at least one live birth and reported the birthweight of their first live born child (N=165,184). Supplementary Table 1 summarises how each variable used here were measured in UKB and coded in our example. We also randomly split those genetically unrelated women of European descent into two groups (Supplementary Figure 1) to obtain SNP specific summary statistics for two-sample MR in this illustrative example. We selected the SNPs for these analyses from the published GWAS of insomnia^(37)^, height,^(38)^ BMI,^(39)^ age at first live birth^(40)^ and education^(41)^ in women and from the previous GWAS of frequency of alcohol intake and ever smoking in UKB men and women.^(42)^ We obtained both SNP-exposure and SNP-outcome results from both of the random sub-samples and the pooled results from analyses in which sample 1 was used for SNP-exposure and sample 2 for SNP-outcome with those in which sample 1 was used for SNP-outcome and sample 2 for SNP-exposure (Supplementary Methods).

#### Exploring the role of population stratification

Each additional allele in the insomnia PRS was associated with a variation of −0.004 (95% confidence interval [CI]: −0.007, −0.001) year in age at recruitment, −3.7×10^−7^ (95% CI: −6.7×10^−7^, −6.4×10^−8^) metre (M) in longitude of birthplace and 2.1×10^−7^ (95% CI: 5.7×10^−8^, 3.5×10^−7^) M in latitude of birthplace. There was evidence of some variation in the mean PRS across 22 UKB assessment centres (Supplementary Figure 2; P-value = 9.2×10^−8^). After adjusting for genetic array, top 40 genetic principal components, participants’ age, birthplace and assessment centre, associations of the PRS with height, BMI, education, frequency of alcohol consumption and ever smoking were not attenuated (Figure 1) suggesting these associations are unlikely to be driven by PS. The association of the insomnia PRS with age at first live birth was slightly attenuated to the null (Figure 1), suggesting some of this may be due to confounding by PS. However, we obtained similar estimates in the MR analyses before and after adjustment for sources of PS (Supplementary Table 2).

#### Distinguishing vertical from horizontal pleiotropy and accounting for horizontal pleiotropy

We searched for GWAS to identify genetic IVs for each of the six non-exposure traits that were conducted in samples independent of UKB and had results presented in women only. However, we were unable to find such GWAS of frequency of alcohol intake or ever smoking. Full details of the selected SNPs are provided in Supplementary Data 1. We found that height, BMI and frequency of alcohol consumption were more likely to reflect vertical pleiotropy or not associated with birthweight (Figure 2), suggesting the associations of the PRS with them were unlikely to introduce bias. These findings have some consistency with previous MR studies.^(18, 43-45)^ However, age at first live birth, education and ever smoking were plausible sources of horizontal pleiotropy (Figure 2). After adjusting for these in MVMR, the effect estimates of insomnia on birthweight attenuated towards the null compared to univariable MR (Figure 3), though results are imprecise.

**Figure 2.**
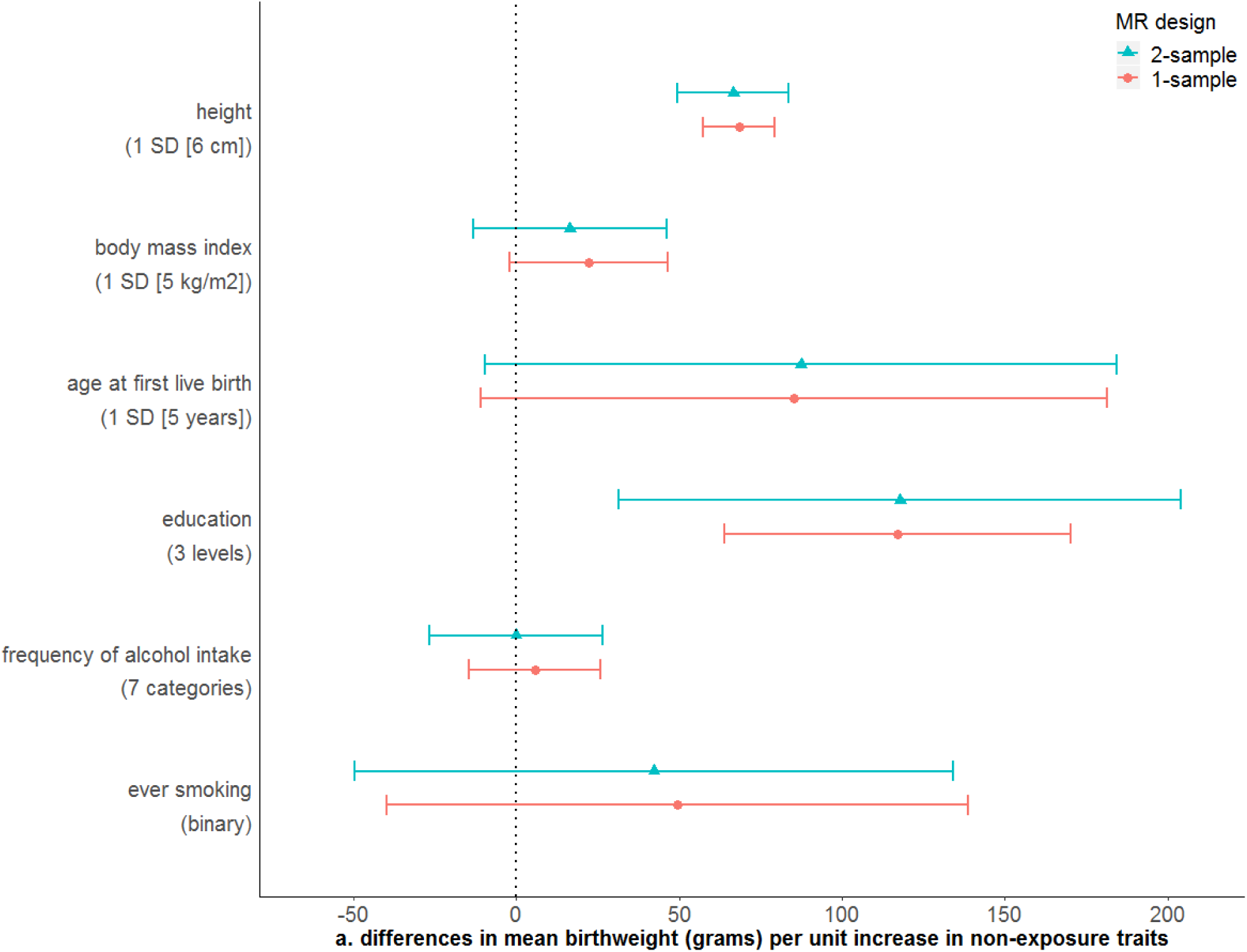
Mendelian randomization estimates for (a) non-exposure traits-birthweight (W-Y) effects, (b) non-exposure traits-insomnia (W-X) effects, and (c) insomnia-non-exposure traits (X-W) effects. “Usually” having insomnia is coded as 1, while “sometimes/rarely/never” having insomnia is coded as 0 (Supplementary Table 1).

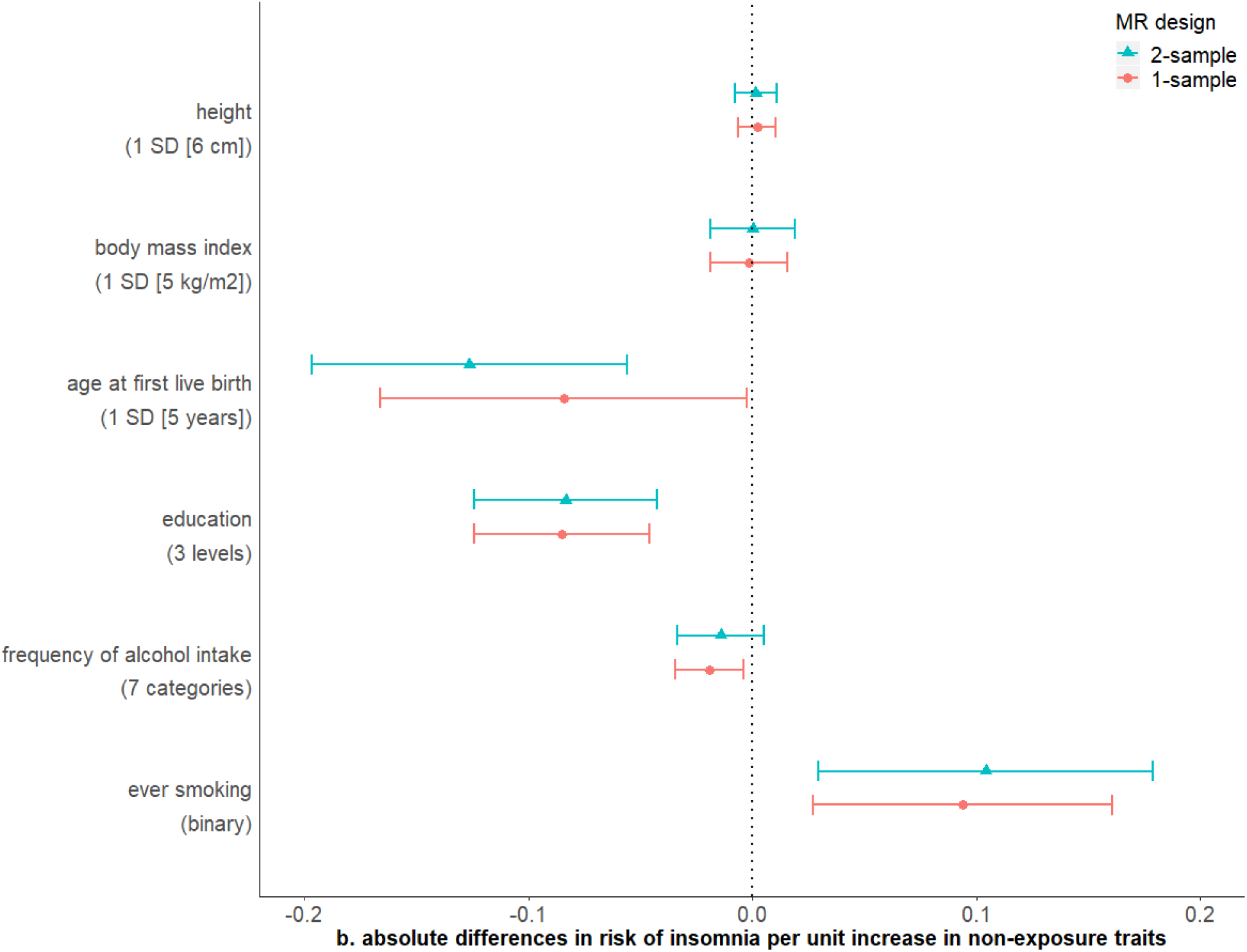

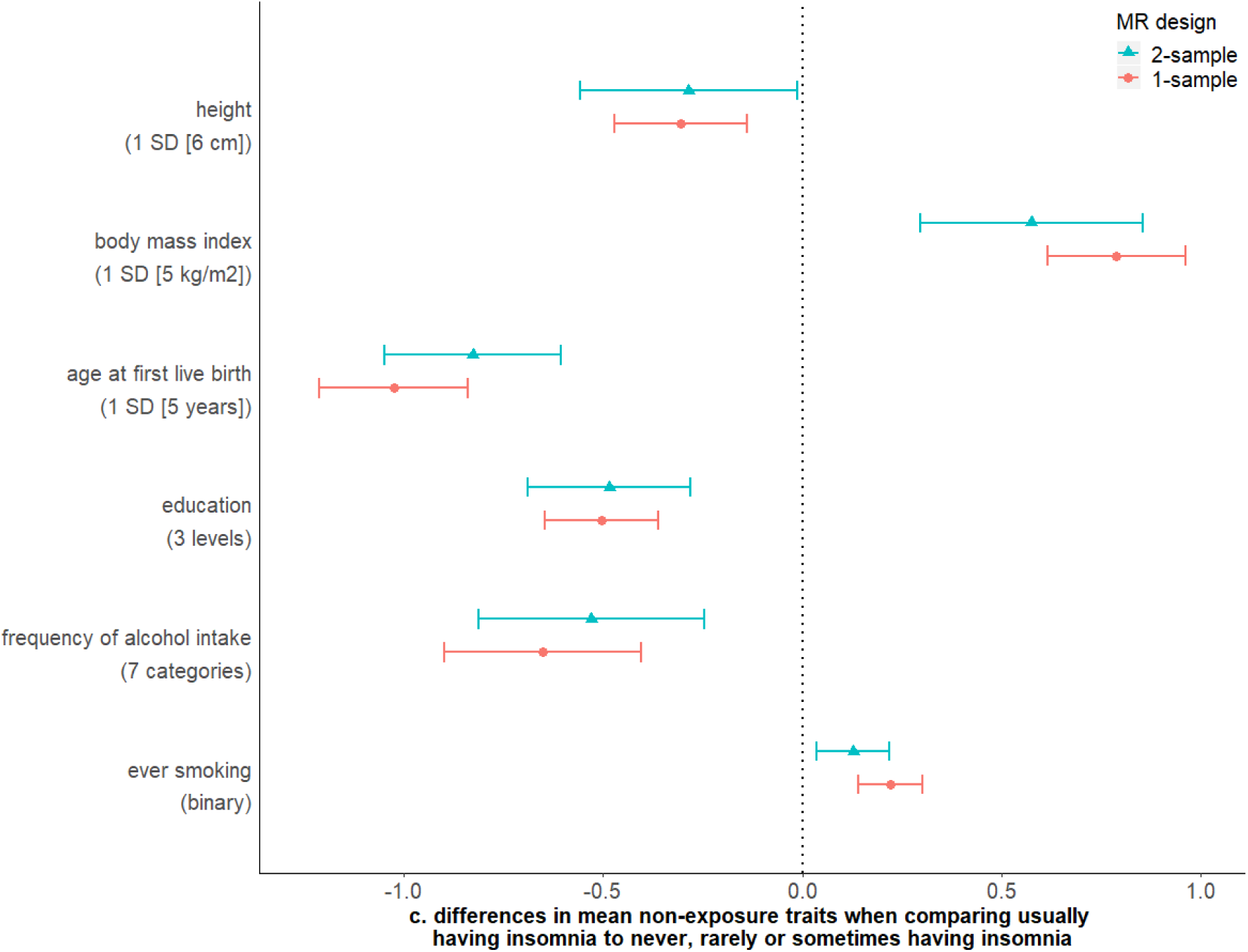

**Figure 3.**
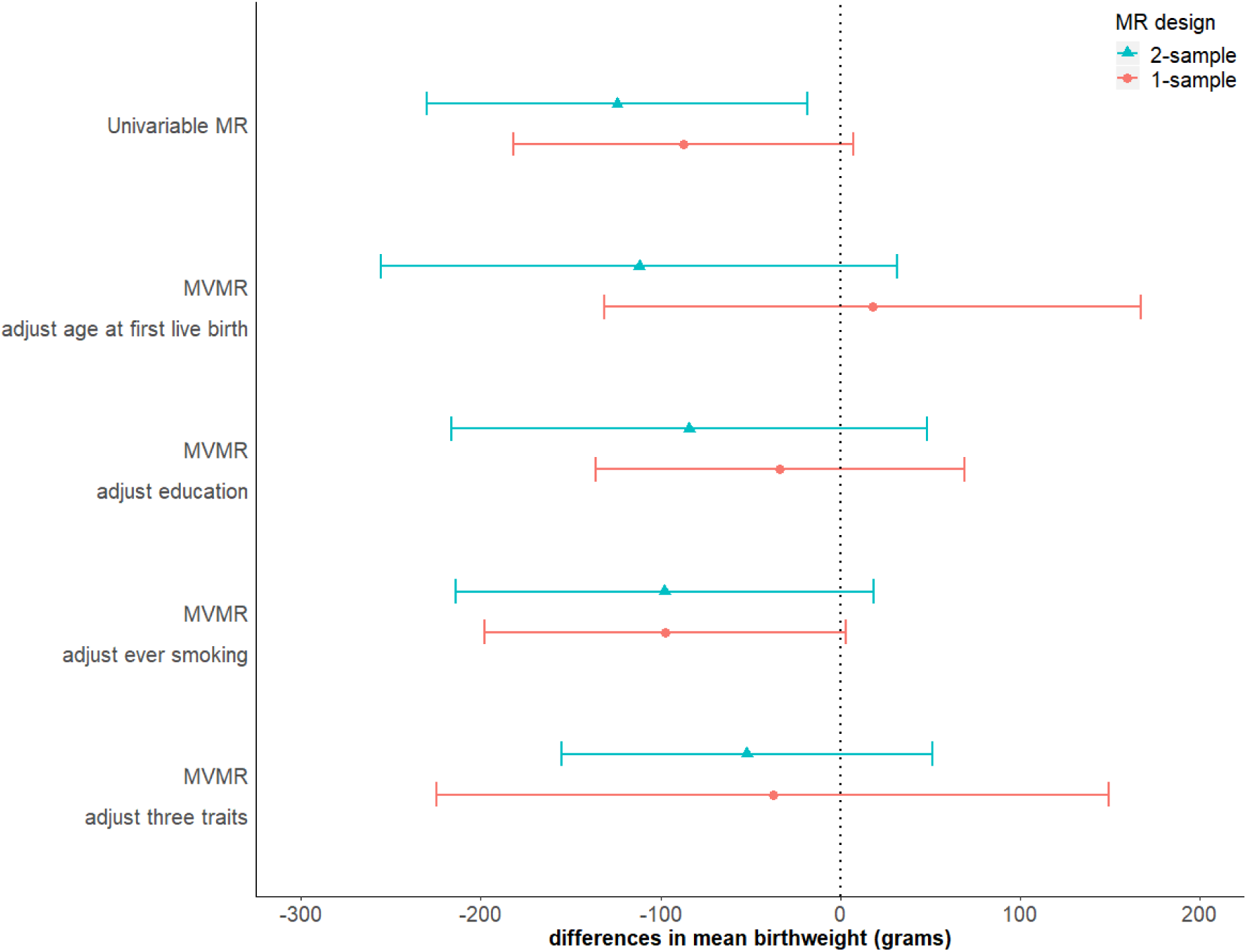
Multivariable Mendelian randomization (MVMR) estimates for the effect of maternal insomnia on offspring birthweight. Estimates are differences in mean birthweight when comparing reporting usually experiencing insomnia to never, rarely or sometimes experiencing it with and without adjustment for potential horizontal pleiotropy via maternal age at first birth, education and ever smoking.

In sensitivity analysis in one-sample MR, sisVIVE (full results in Supplementary Data 2) suggested that the association of insomnia with birthweight was greater than seen with univariable TSLS (−87 [95% CI: −182, 7] grams). In the two sample MR results from all sensitivity analyses were directionally consistent with the main IVW estimate, though for several the CIs were very wide; IVW, MR-PRESSO and MR-TRYX supported an inverse association of maternal insomnia with offspring birthweight with CIs that did not include the null (Figure 4). The MR-Egger intercept suggested little evidence of unbalanced horizontal pleiotropy (p-value = 0.732 for dataset A on B and 0.763 for B on A; full results in Supplementary Figure 3). Whilst between SNP heterogeneity was less when MR-TRYX was used (in comparison to the IVW analyses) the point estimates were very similar between it and IVW (Figure 4 and Supplementary Figure 4).

**Figure 4.**
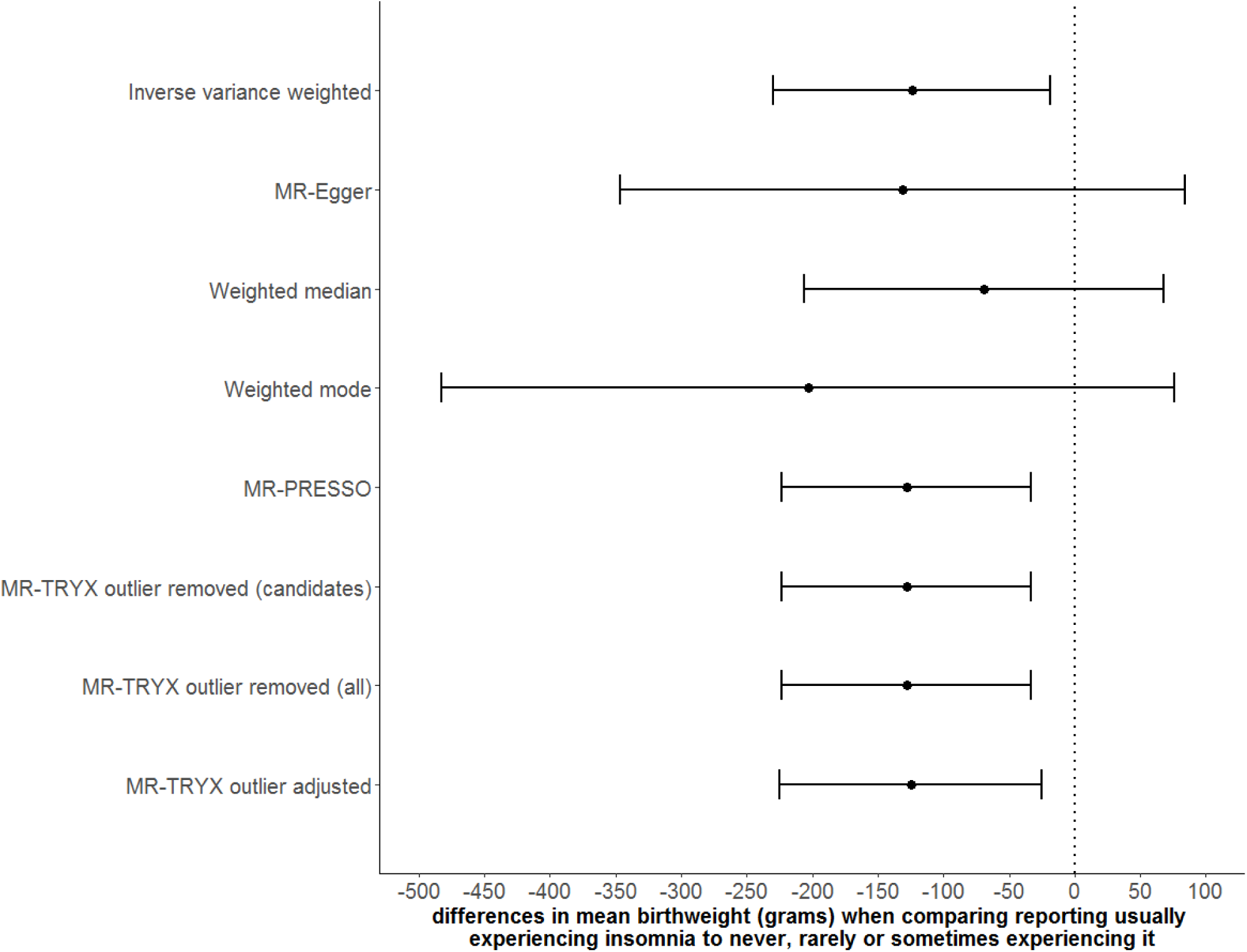
Sensitivity analyses for the effect of maternal insomnia on offspring birthweight using two-sample Mendelian randomization (MR)

#### Identifying more potential sources of violation of MR assumptions using a phenome-wide approach

In this motivating example we only explored the six traits that we had *a priori* selected for checking, based on prior knowledge that these were key risk factors for variation in birthweight. However, there may be value in exploring a wider range of potential violating paths (recommendation 1). Therefore, we undertook a comprehensive search for previously identified genome-wide significant associations of the 80 SNPs in the insomnia PRS using Phenoscanner.^(15)^ This amounted to 478 associations that included 42 SNPs, among which 34 SNPs were associated with at least one trait apart from sleep (full results in Supplementary Data 3). We did not examine these further but discuss in the discussion section the pros and cons of different approaches to identifying genetic IVs associations with non-exposure traits.

#### Exploring reverse causality

Whilst temporally it is hard to conceive of birthweight influencing maternal insomnia, birthweight is a proxy for fetal growth, which could influence maternal insomnia symptoms. To explore this possibility, we would require offspring (fetal) genetic variants that are robustly related to their fetal growth. Whilst there are no GWAS currently of maternal or fetal contribution to fetal growth (e.g. assessed by repeat ultrasound scan) there are GWAS of own (i.e. fetal) genetic variants in relation to own birthweight.^(46)^ However, we do not have genome-wide data in maternal-offspring pairs in UKB and so cannot explore this here.

## Discussion

The possibility that genetic IVs for a specific exposure will associate with many other traits is increasing as GWAS explore a larger number of SNPs in increasing sample sizes. In this paper we have described different scenarios that could result in such associations and methods for exploring where these may cause bias. Beyond confounding by PS, a key concern is attempting to differentiate vertical from horizontal pleiotropy and using methods to explore and reduce bias from horizontal pleiotropy. We provide a set of recommendations and demonstrate their use in an applied example exploring the effect of maternal insomnia on birthweight.

This paper brings attention to the pros and cons of hypothesis-driven versus comprehensive approaches to exploring IV validity. Our motivating example used researchers’ knowledge to decide which non-exposure traits to explore genetic IVs associations with. Specifically, we chose six observed traits in UKB that we considered plausible causes of offspring birthweight, and as our analyses in this example shows they reflect plausible horizontal or vertical pleiotropic paths. However, we have to rely on sensitivity analyses (see Table 2) to control for horizontal pleiotropy via unexplored traits. This approach is efficient but cannot identify the nature of any unexplored violation of instrument validity. Sensitivity analyses will identify whether results are likely to be biased by unbalanced horizontal pleiotropy but if one wanted to explore specific known horizontal or vertical (mediating) pleiotropy this approach would potentially miss some key paths. A further limitation is that researchers’ knowledge is likely to vary between different researcher groups. An alternative to *a priori* selecting a defined set of potential pleiotropic traits, is to use a Phenome-wide search to systematically explore any possible non-exposure traits that associate with our genetic IVs. This approach has the advantage that it is not limited by researchers’ own knowledge and the variation in this between research groups. Although automated systems for rapid phenome-wide associations now make a more extensive and systematic approach possible,^(47-49)^ there are challenges in applying our recommendations 3-5 to a possible large number of traits.

Of particular importance, when multiple different potential traits (exposure of interest and non-exposure traits) and the relationship between them is being considered differing measurement error in each trait may affect the results obtained. In MR and MVMR differing measurement error in different traits gives the same effect as differing power in each trait and will lead to the effects of traits measured with more error being less precisely estimated than the effects of those measured with less error. However, Steiger filtering assumes that each trait is measured with the same error and can give misleading results for the causal direction between two traits when the true causal exposure is measured with more error than the true outcome. In our example self-report of insomnia is likely to be measured with more error than several of the non-exposure traits considered, in particular maternal height, BMI, age at first birth and education. For these traits Stieger filtering may misleadingly suggest that the direction of effect is from these traits to insomnia due to the imprecision in the measurement of insomnia. These issues are relevant to both the use of prior knowledge to select specific traits to explore as possible pleiotropic paths and to a more comprehensive and systematic phenome-wide scan approach. However, with the latter there are many more non-exposure traits where these problems are likely to arise. In our illustrative example the phenome-wide scan approach identified 478 non-exposure traits associated with one or more of the 80 insomnia SNPs used in our genetic IV (i.e. 80-fold the six explored on the basis of prior knowledge). On-the-one hand this suggests we might have missed some key specific pleiotropic paths, on the other, even with the large sample size used in our example the potential for uninterpretable imprecise results and possible misleading results is increased with the much larger number from the phenome-wide scan.

Finally, the automated phenome-wide approach is dependent on the nature and quality of the studies included in the searching tools (e.g. PhenoScanner^(15)^ and GWAS Catalog^(16)^) and whilst they are likely to identify more specific pleiotropic paths than knowledge based approaches, they may still miss some important paths. Whether researchers decide to focus solely on a limited set of traits that are known through prior knowledge to influence outcome and could be on a horizontal pleiotropic path or undertake a phenome-wide approach will depend on the specific research question, including whether that includes an interest in understanding the nature of horizontal pleiotropic paths or mediation (vertical pleiotropy). It will also depend on available data. A combination of both could be undertaken with some a prior decision to select a fixed number of the non-exposure traits identified by the searching tool.

Our study provides guidance for further MR studies where genetic IVs were associated with multiple traits. It may also be relevant to studies using non-genetic IVs (e.g. healthcare practitioner preference^(50)^ or randomization in a randomized control trials^(51)^). In addition to the approaches outlined here to the situation of identifying that genetic IVs are related to multiple non-exposure traits, we would recommend triangulating MR results with other methods that have different key sources of bias to estimate causal effects.^(52)^ If results are consistent across such different methods that increases confidence in the result, even in the presence of remaining concerns about genetic IV validity.

## Data Availability

The data reported in this paper are available by applying directly to the UK Biobank.

https://www.ukbiobank.ac.uk

## Funding

This work was supported by the University of Bristol and UK Medical Research Council [grant numbers, MM_UU_00011/1, MM_UU_00011/2, MM_UU_00011/3 and MM_UU_00011/6], the US National Institute for Health (R01 DK10324),the European Research Council via Advanced Grant 669545, the British Heart Foundation (AA/18/7/34219) and the NIHR Biomedical Research Centre at University Hospitals Bristol NHS Foundation Trust and the University of Bristol. Q.Y. is funded by a China Scholarship Council PhD Scholarship (CSC201808060273). M.C.B. is funded by a UK Medical Research Council Skills Development Fellowship (MR/P014054/1). D.A.L. is an NIHR Senior Investigator (NF-0616-10102). The funders had no influence on the study design, data collection and analysis, decision to publish, or preparation of the manuscript.

## Acknowledgements

This research was conducted using the UK Biobank Resource under Application Number 23938. The UK Biobank received ethical approval from the research ethics committee (REC reference for UK Biobank 11/NW/0382) and participants provided written informed consent.

## Competing interests

D.A.L. reports receiving research support from Medtronic and Roche Diagnostics for research outside the submitted work. All other authors declare no competing interests.

## Notes

### Author Declarations

All relevant ethical guidelines have been followed and any necessary IRB and/or ethics committee approvals have been obtained.

Any clinical trials involved have been registered with an ICMJE-approved registry such as ClinicalTrials.gov and the trial ID is included in the manuscript.

